# Accounting for the Family Physician Workforce in Newfoundland and Labrador: A Stock and Flow Analysis, 2014–2024

**DOI:** 10.64898/2026.01.10.26343845

**Authors:** Maisam Najafizada, Arista Marthyman, Elhamy Samak, Kris Aubrey-Bassler

## Abstract

**Introduction:** Newfoundland and Labrador (NL) faces persistent difficulty attaching its residents to primary care. We undertook a stock and flow analysis to represent how inflows and outflows of family physicians (FPs) shape effective capacity and to provide a reconciled estimate of FP supply for 2024. This approach clarifies drivers of change, exposes intervention points, and supports timely planning.

**Methods:** We assembled a multi-year headcount series and linked it to CIHI’s “entering/leaving direct care” flows, harmonizing definitions and time frames across sources. We compared observed year-to-year stock change with net flows to identify timing and classification gaps. Stakeholder consultations informed key parameters (graduates and retention, internationally trained entrants, migration, retirement, and scope shift). Because confirmations are released with a lag, we produced a reconciled 2024 estimate using the CIHI headcount as baseline and these validated inputs.

**Findings:** FP headcount changed from 680 (2014) to 666 (2023) (–2.1%) after peaking at 728 (2017); the ratio fell from 129 to 124 per 100,000 population. The workforce became more urban (rural 255→203; urban 424→460) and more Canada-trained (417→466) while foreign trained decreased (261→199). Net interprovincial migration averaged –24/year, with pronounced losses in 2019 (–57) and 2022 (–42). CIHI entry–exit data point to marked volatility in the FP workforce: entries/exits were 110/96 (2019), 62/88 (2020), and 71/117 (2021), with residuals versus stock change indicating definitional/timing differences. The 2024 reconciliation yielded ≈658 FPs (net –8.5 from 2023), ≈507 FTE at 0.77 FTE/head, and ≈122 per 100,000 population.

**Conclusion:** Inflows from local graduates and IMGs did not fully offset exits from migration, retirement, and scope/burnout in 2024. Recruitment alone is unlikely to close access gaps; retention-first strategies, scaleup of team-based care with role optimization, targeted rural supports, and routine monitoring of flows are needed to stabilize and grow effective primary care capacity in NL.

## Introduction

Access to comprehensive, continuous primary care is a cornerstone of high performing health systems, with strong associations between family physician (FP) supply and better population health outcomes, lower hospitalization rates, and more equitable care [1]. In Newfoundland and Labrador (NL), persistent difficulty accessing a regular FP has become a defining system challenge. Recent polling suggests that roughly 30% of Newfoundlanders and Labradorians—approximately 163,000 people—report not having a regular family doctor [2]. This access problem is amplified by the province’s unique geography and demography: a vast territory with low population density (1.4 persons/km²) and many small, rural communities, which complicates equitable distribution of the workforce and continuity of care [3].

Recognizing the urgency, Health Accord NL released a 10-year transformation blueprint in 2022 that elevated primary care reform as a central lever for system sustainability, including the establishment of team-based Family Care Teams (FCTs) to expand access and continuity [4]. In parallel, the province consolidated its health system into a single authority—NL Health Services—in April 2023 to streamline planning and delivery and to enable provincewide primary care redesign [5]. Since 2022, government and NL Health Services have announced multiple FCTs and reported substantial progress in connecting residents to these teams [5, 6].

At the national level, the Canadian Institute for Health Information (CIHI) documents important trends shaping FP availability. While the number of FPs in direct care increased over the past decade, overall growth has slowed in recent years, and effective supply should be assessed in terms of fulltime equivalents (FTEs) rather than headcounts alone [7]. Moreover, CIHI emphasizes that “effective” FP supply per 100,000 population is lower than simple headcount ratios, underscoring the importance of workload and practice patterns in workforce planning [7]. These considerations are particularly salient for NL, where aging populations, rural distribution, and interprovincial mobility can produce local shortages even when provincial totals appear stable.

Stock and flow models—long used in Organisation for Economic Co-operation and Development (OECD) countries for health human resources planning—make explicit the dynamic processes that change workforce capacity over time: training and recruitment inflows (e.g., new graduates, international medical graduates, interprovincial in-migration), internal flows (e.g., changes in FTE, scope, or practice model), and outflows (e.g., retirement, outmigration, career change) [8, 9]. Canadian scholars have adapted and extended these methods to simulate supply and requirements under different policy scenarios, highlighting how planning based on needs and service models can improve alignment between capacity and population health needs [10, 11]. A province specific stock and flow analysis for NL can therefore quantify current capacity, illuminate the drivers of FP shortages, and test the impact of realistic levers—such as enhanced retention, expanded FCT uptake, role optimization for nurse practitioners, and recruitment pathways—on medium and long-term access to primary care.

Despite active reforms, publicly accessible, integrated analyses that combine administrative data from multiple sources with stakeholder insights to estimate NL’s FP “stocks” and “flows” remain limited. Given the scale of ongoing change—FCT implementation, the creation of a single health authority, and evolving recruitment/retention strategies—decision makers require up-to-date, transparent modelling to assess whether policy trajectories are sufficient to close access gaps and to identify where course corrections are needed [4, 7].

## Objective

To conduct a provincewide stock and flow analysis of the FP workforce in Newfoundland and Labrador using publicly available data and consultations with key stakeholders, in order to (a) describe current FP supply patterns; and (b) quantify current inflows, internal flows, and outflows.

### Research Questions

1. **Current capacity and distribution.** What is the current FP stock in NL by headcount and effective FTE, and how is it distributed across zones, rural/urban settings and practice models (e.g., FCT-affiliated, fee-for-service, salaried)?
2. **Inflow and outflow dynamics.** What are the dynamics of inflows (new graduates, international medical graduates, interprovincial in-migration), internal flows (changes in FTE, scope, or practice model), and outflows (retirement, outmigration, career shift)?

## Methodology

### Study design and setting

We conducted a retrospective stock and flow analysis of NL’s FP workforce. Originating in system dynamics, stock-and-flow modeling tracks how accumulations change through inflows and outflows; this approach is now common in health resources for human planning to examine training pipelines, migration, and retirement [8, 9]. Stocks were presented as both headcount by calendar year and FTE conversion using a derivation from CIHI’s head count to FTE calculations [12]. Flows were determined from publicly available data on FP entries and exits to/from direct patient care in primary healthcare settings. Analyses were performed at the provincial level, with descriptive subgrouping by gender, rural/urban location, and place of MD graduation where reported. Historical stocks were compiled for 2014–2023; CIHI “entering/leaving direct care” flows were compiled for 2019–2021, and a 2024 stock and flow reconciliation was constructed and validated with system stakeholders.

### Data sources and case definitions

Primary data were drawn from the Canadian Institute for Health Information (CIHI) physician supply series and direct care flow tables; Statistics Canada intercensal population estimates for denominators; and provincial/organizational sources for 2024 parameters (Faculty of Medicine reports [13–20], PGME records, NLMA postings, CPSNL licensure information, and PRANL confirmations). “Family physician” followed CIHI’s *family medicine/general practice* category. Population denominators and rural/urban, gender, and training origin breakdowns correspond to the fields in the assembled NL dataset.

#### Time frame and datasets

- **Stocks (2014–2023):** Annual NL FP headcounts with covariates (male/female; rural/urban; Canada vs foreign trained), accompanying population counts, and computed physician-to-population ratios.
- **Flows (2019–2021):** CIHI counts of entries and exits to/from direct care for NL
- **2024 Reconciliation Parameters:** 2023 baseline stock; head count → FTE conversion; MUN Family Medicine graduates with retention percentage; IMG inflow; net interprovincial migration; moving abroad; return from abroad; retirement rate; burnout/scope shift exit.

#### Variables and measures

- **Stock (N):** Year-end FP headcount.
- **Entries (E) / Exits (X):** CIHI direct care flows (2019–2021) by fiscal year; for 2024, components parameterized from provincial/organizational sources.
- **Effective capacity (FTE):** Headcount × **0.77** (we used 10 years average using CIHI NPDB payment-to-FTE ratios).
- **Retirement rate (1.8%).** We parameterized retirement at **1.8% annually**, using **Alberta’s higher-end estimate (∼1.2–1.8%)** as a conservative proxy for NL. Choosing the upper bound minimizes the risk of undercounting exits in the 2024 reconciliation.
- **Burnout/scope-shift exits**. We estimated 1.4% by: (1) comparing CIHI’s 2019–2021 entry/exit figures with the headcount change we observed, and (2) adjusting the rate so it also fit what stakeholders reported in 2024
- **Density:** FPs per **100,000** residents using Statistics Canada annual estimates.

### Data extraction, cleaning, and harmonization

We imported the CIHI tables and provincial datasets and combined them into one long-format dataset with standard variable names and year labels (stocks = total headcount for 2014–2024; flows = entries/exits for 2019–2021). We then added annual population estimates so we could calculate physicians per 100,000 residents each year. For 2019–2021, we compared the actual year-to-year change in headcount ((N_t - N_{t-1})) with the net of entries and exits ((E_t - X_t)). Any difference between these two numbers was recorded as a residual, which we attribute to definition or timing mismatches between headcount totals and CIHI’s “direct care” flow series. We used these residuals to guide—but not override—our reconciliation.

### Stakeholder consultations

After assembling the historical headcounts and CIHI flow data, we shared a brief evidence pack and held targeted consultations with stakeholders from Family Medicine and PGME (Faculty of Medicine), the NLMA, and NL Health Services. Using a draft 2024 worksheet, stakeholders checked key components—such as the number of MUN Family Medicine graduates and their retention, IMG licence counts, net interprovincial losses, and estimates for retirement and scope shift. They also provided context to help interpret the results. We only made changes to the 2024 reconciliation when they were backed by documented sources or agreed to by multiple stakeholders.

### Analysis

We first constructed a descriptive time series for 2014–2024, reporting annual family physician (FP) stocks and describing composition (gender and place of MD graduation), rural/urban distribution, and physician-to-population density. We then analyzed flows for 2019–2021 by tabulating CIHI “entering” and “leaving” direct care counts, computing gross turnover and net flows, and contrasting these with observed year-over-year stock changes to assess internal coherence and quantify reconciliation residuals. Finally, we produced a 2024 stock and flow table using the accounting identity N=666 and parameterizing **entries** (inflows) as retained Memorial University Family Medicine graduates, new IMG licences, and returns from abroad, and **exits** (outflows) as net interprovincial migration, departures abroad, retirements, and burnout/scope shift exits.

Throughout the process, we maintained a simple hierarchy of evidence (CIHI and official public records > organizational reports > multistakeholder consensus) for parameter selection and change control. All analyses used aggregate, publicly available information and organization level inputs; no patient level or personally identifiable data were involved, and thus institutional ethics review was not required.

## Findings

### 1) Historical stock of family physicians (2014–2023)

From 2014 to 2023, NL FP headcount decreased modestly from 680 to 666 (–2.1%). The series peaked at 728 in 2017 and then declined by 62 physicians to 2023 (–8.5%). Over the same period, the provincial population rose slightly (≈528,000 to 538,605), and the physiciansper100,000 ratio fell from 129 (2014) to 124 (2023), down 5 points from 2014 and 13 points from the 2017 high (137).

**Table 1.** NL Family Physician Counts and Characteristics 2014-2023.

The composition and distribution of this stock show insightful findings. Over the decade, the workforce became more gender-balanced and more urban-concentrated:

- **Gender:** Female physicians rose from 40.4% (275/680) in 2014 to 47.9% (319/666) in 2023, a +7.5point shift; male headcount fell 14.1% (404→347), while female headcount rose 16.0% (275→319).
- **Geography:** Rural headcount declined 20.4% (255→203), while urban headcount increased 8.5% (424→460). Rural share fell from 37.5% to 30.5% (–7.0 points).
- **Training origin:** Canada-trained physicians increased 11.8% (417→466) as foreign-trained decreased 23.8% (261→199). The Canada-trained share rose from 61.3% (2014) to 70.0% (2023) (≈+8.6 points).

**Figure 1.**
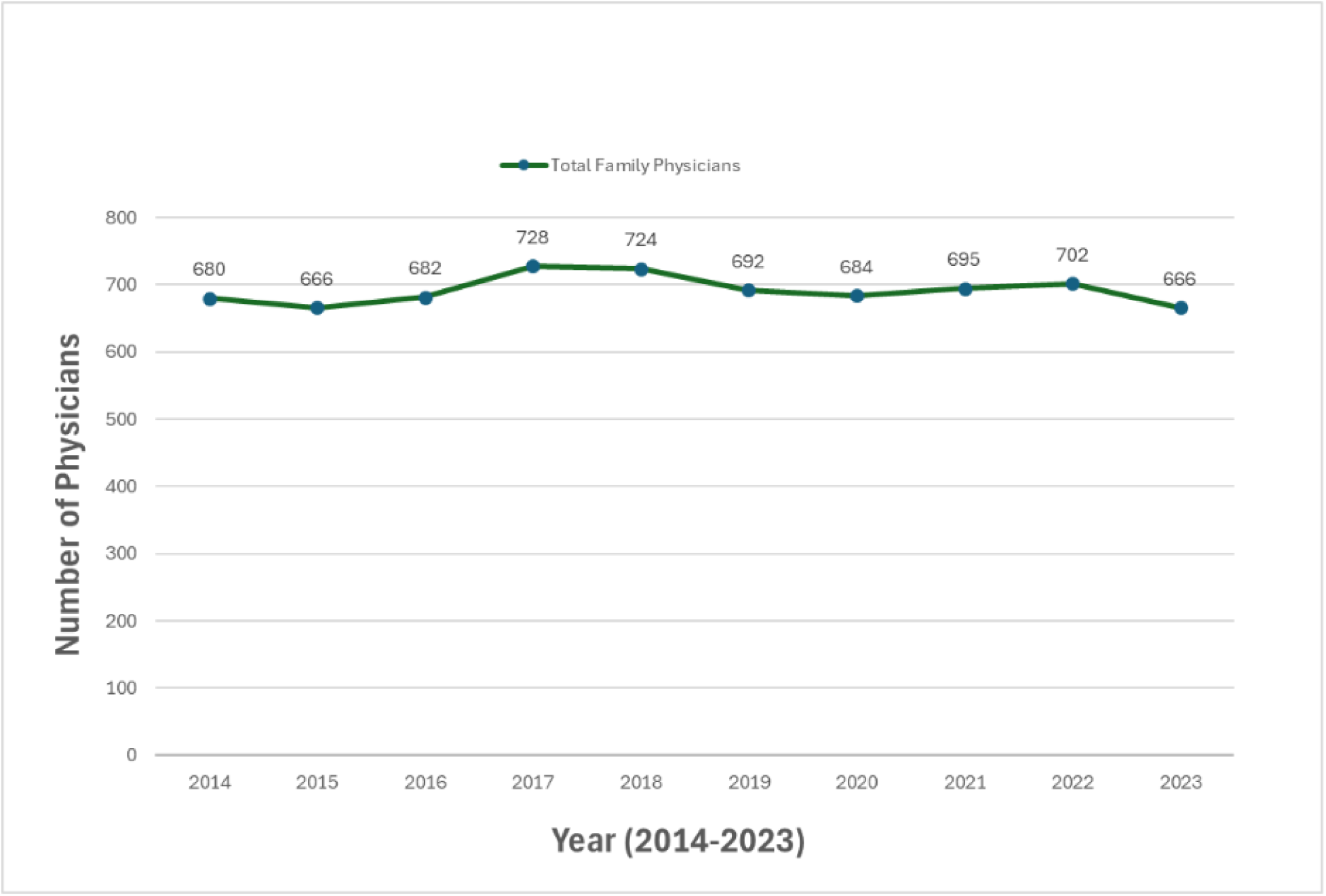
Number of NL Family Physicians (Head count) in Primary Care by Year:

**Figure 2.**
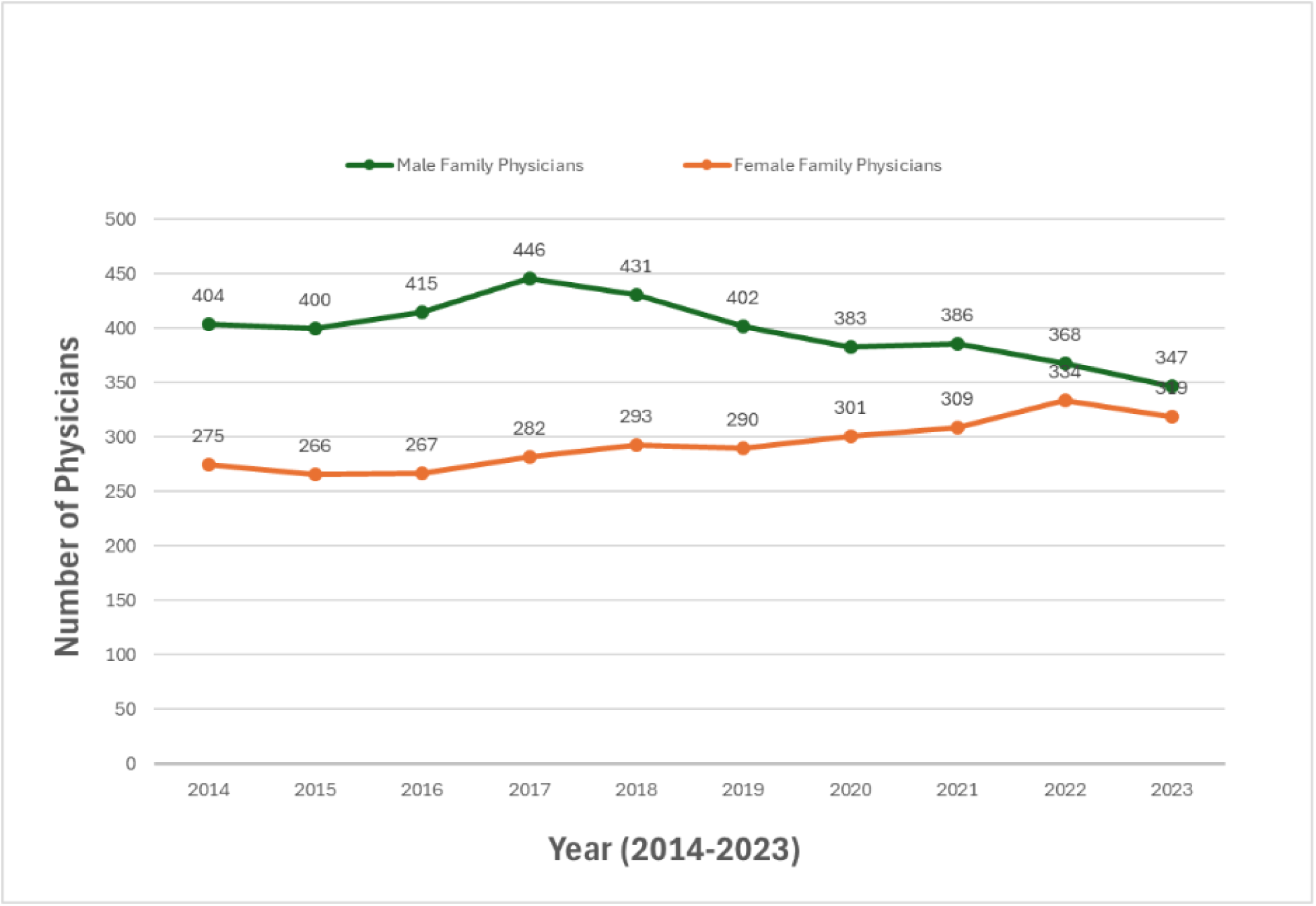
Number of NL Family Physicians in Primary Care by Sex and Year:

**Figure 3.**
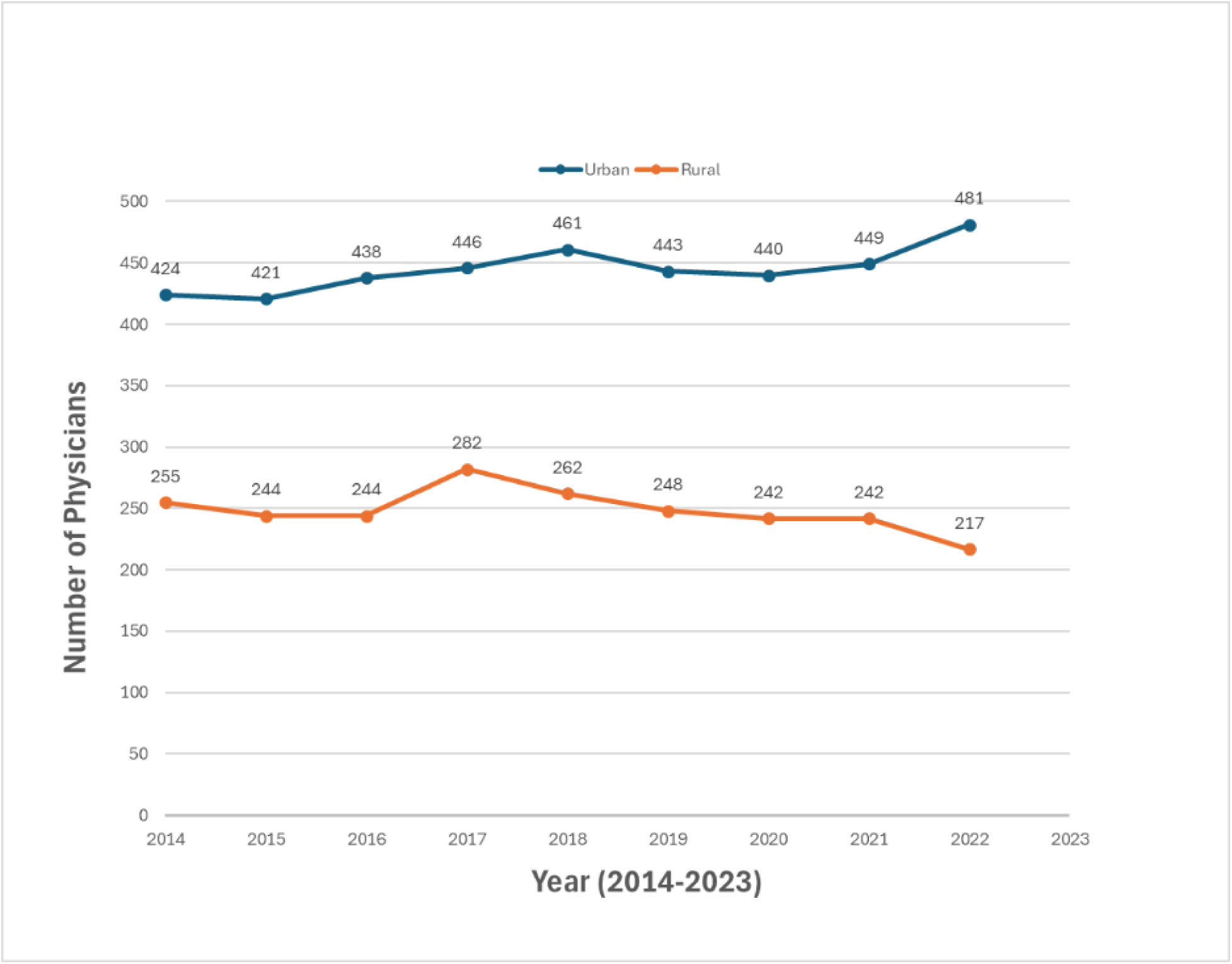
Stock Rural vs Urban Distribution by Year:

**Figure 4.**
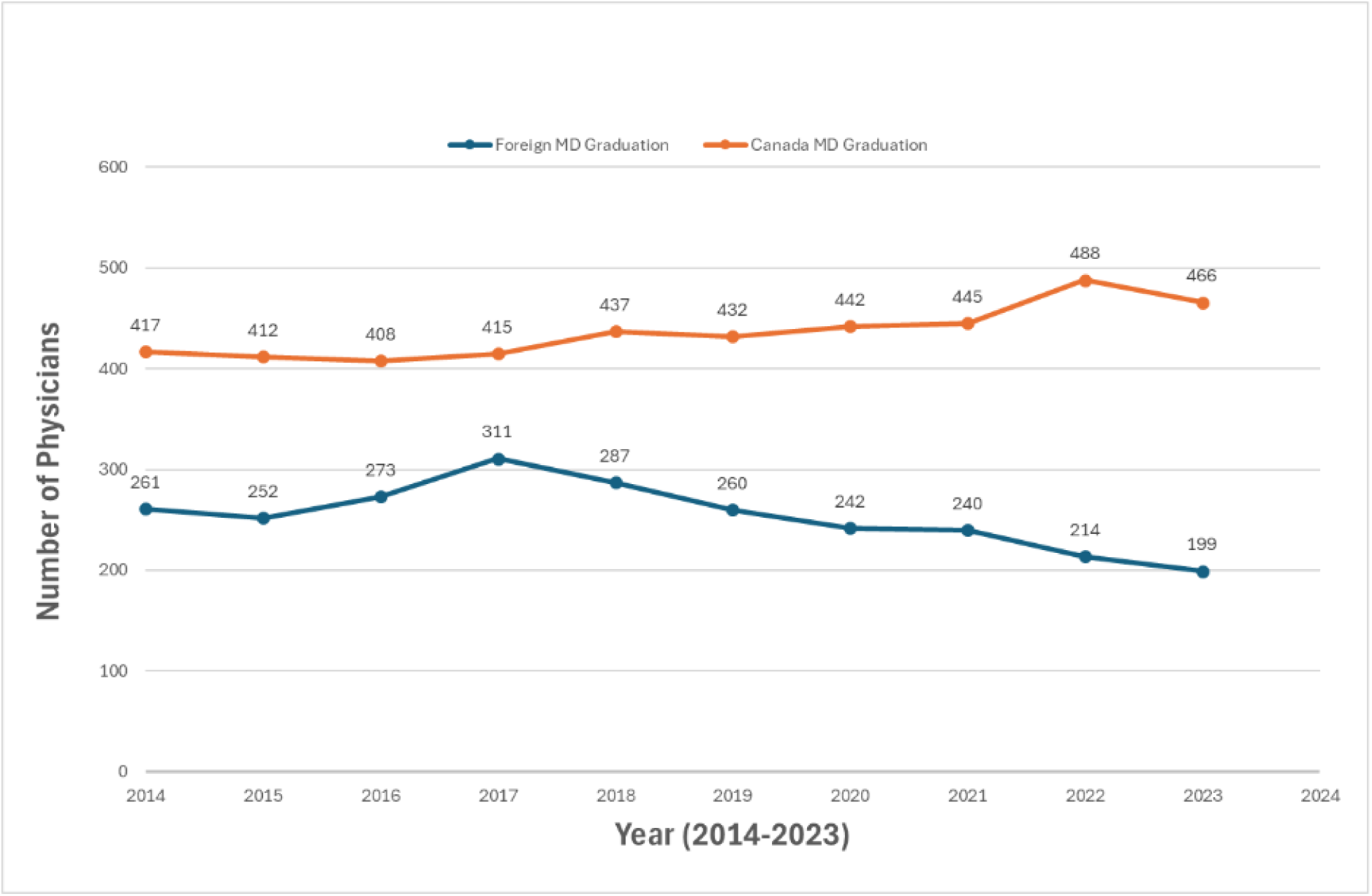
NL Family Physician Numbers by Location of MD Graduation and Year.

One of the most significant influences on family physician stock has been interprovincial migration, which was a net loss averaging –24 physicians/year from 2014-2023 (range: –2 to – 57). Large decreases in stock occurred in 2019 (–57) and 2022 (–42), with partial improvement to –25 in 2023. These net losses are material given the province’s total numbers of family physicians.

### 2) Entries and exits (CIHI direct care flows), 2019–2021

CIHI’s “entering/leaving direct care” series shows substantial **gross turnover:**

**Table 2.** NL Family Physician Entries and Exits, 2019-2021. *Rates use prior year end of year stock as denominator. †“Reconciliation residual” = (observed change in stock) – (Entries – Exits); positive values indicate growth not explained by reported flows, negative values indicate decline not explained by reported flows. The residuals likely reflect definitional and timing differences between headcount stock and “direct-care” flow series.

Despite a net positive flow in 2019 (+14), the stock fell (–32), yielding a –46 residual; conversely, in 2021, net flows were negative (–46) while stock rose (+11), producing a +57 residual. Since the flow numbers and the headcount totals do not reconcile tear-to-year, that mismatch likely comes from differences in data collection, definitions (e.g., “direct care” vs. total headcount) and timing (fiscal vs. calendar). Because of these gaps, we used a reconciliation step for 2024 that lets the documented flows inform the result, while adjusting for those known definition/timing differences.

### 3) 2024 stock and flow table

We find that the 2024 stock is 666 family physicians (≈513 FTE at 0.77). Inflows are mainly new IMG licences (∼20), MUN FM residency retention (∼18 at 56%), and a small return-from-abroad (∼1). Outflows include retirement (∼12), burnout/scope shift (∼9), physicians moving abroad (∼1), and a large net interprovincial loss (∼25), together indicating a net decline in the stock. (Figure xx; Table xx)

**Figure 5.**
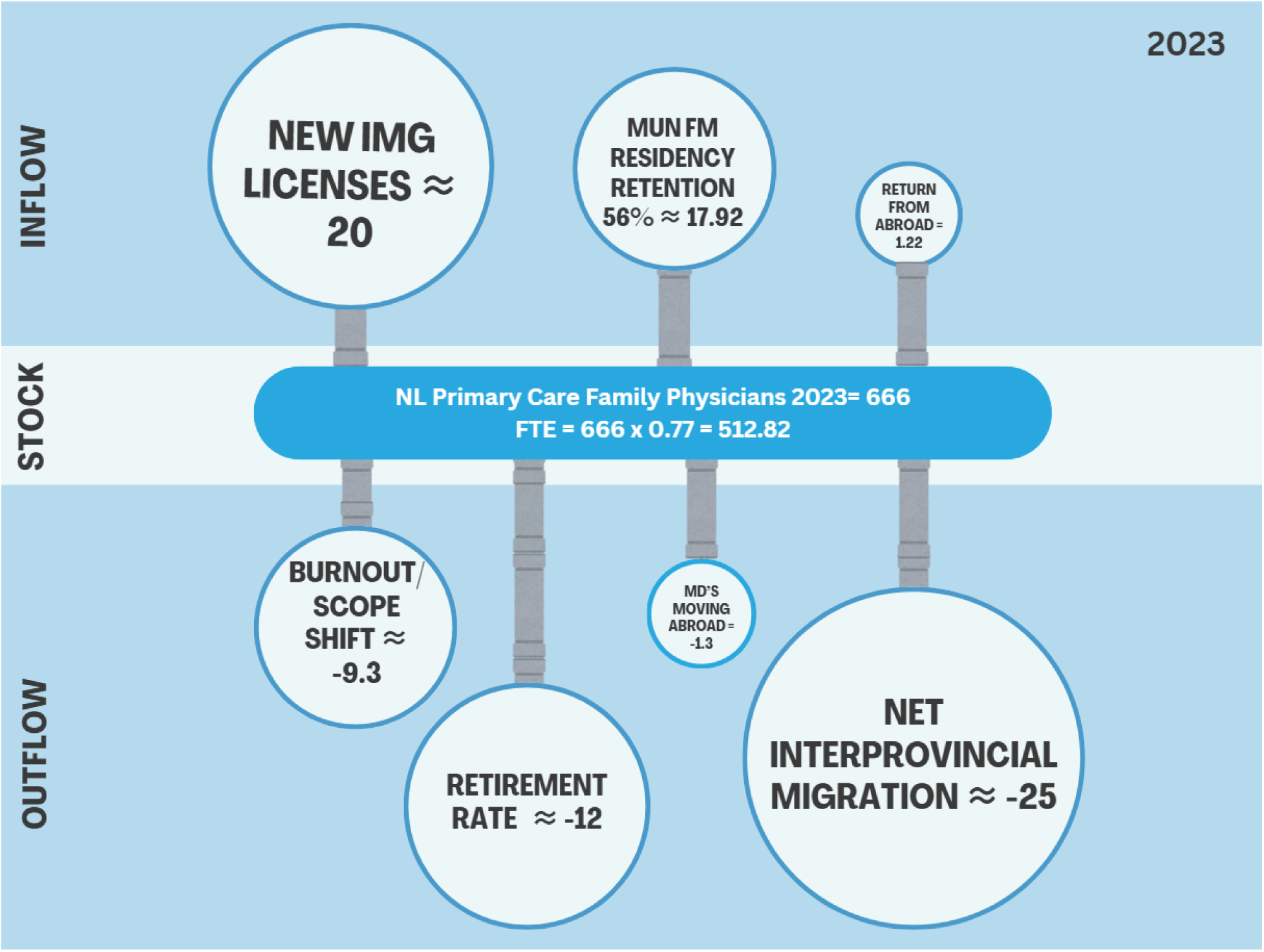
Stock and Flow Diagram of NL Family Physicians in 2023.

**Table 3.** Data Parameters, Definitions and Sources.

Taking CIHI’s reported 2024 family-physician headcount of 666 as the starting stock and then applying parameters validated in our stakeholder consultations (e.g., MUN Family Medicine graduates and retention, IMG inflow, interprovincial migration, and exit rates), this yields an estimated net decline of about 8–9 physicians in 2024, for an end-of-year stock of roughly 658. (Table 4) Using an FTE conversion factor of 0.77 (to translate headcount to full-time equivalents), this corresponds to about 507 effective FTEs. With the 2023 population (538,605) as a stable reference, the implied 2024 density is approximately 122 family physicians per 100,000 residents, down from 124 in 2023.

**Table 4.** 2024 reconciliation table (components).

## Discussion

Over the last decade, NL’s FP stock was broadly flat but volatile, peaking in 2017 (728) and declining to 666 by 2023, or 124 FPs per 100,000—a five point drop from 2014 and 13 points below the 2017 high. The composition of the workforce also shifted: female FPs increased in both absolute number and share, the Canada trained share rose, and the distribution continued to urbanize as rural headcount fell by ∼20%. Net interprovincial migration was negative in most years (averaging ∼–24 per year), and CIHI’s direct care flow series (2019–2021) revealed substantial movement of the workforce, with large reconciliation residuals versus annual stock change. Using stakeholder-validated parameters, our 2024 reconciliation suggests entries did not fully offset exits, yielding an estimated yearend headcount of ≈658 (≈507 FTE at 0.77 FTE/head) and an implied density of ≈122/100,000.

Our NL-specific pattern aligns with Canada-wide signals from CIHI. Nationally, the number of family physicians in direct care increased over the past decade but growth has slowed sharply in recent years; family medicine posted slightly negative growth in 2023 (–0.1%), even as overall physician supply grew 1.4% [24, 25]. CIHI also documents that family physicians are, on average, seeing fewer unique patients per year and report heavy administrative load, and that a nontrivial share is practising outside traditional primary care (e.g., emergency medicine), all of which depresses “effective” primary care capacity even when headcount is stable [26].

On the demand side, access gaps remain large. Statistics Canada’s CCHS indicates that 17% of Canadian adults (≈5.4 million) lacked a regular primary care provider in 2023, and comparative surveys show Canada lagging peer countries on primary care access [7]. NL mirrors—and likely exceeds—these access challenges: an NLMA poll in November 2024 found ∼30% of residents report no regular family doctor (≈163,000 people) [2]. The significant rural distribution of NL’s population exacerbates access gaps beyond the physician headcount challenges. Taken together, our stock and flow results are consistent with the broader Canadian picture of tight supply, high turnover, and erosion of effective capacity.

Two themes from peer countries resonate with our findings. First, across the OECD, overall physician density has risen over the past decade, yet distributional imbalances—especially rural-urban gaps—persist, and growing headcount does not automatically translate into primary care access [27.28]. Second, several high-income systems report GP-specific strain despite rising total doctor numbers. In England, official workforce series show continued pressure in general practice, with long-run declines among GP partners and ongoing workload concerns [29, 30]. In Australia, the federal GP supply-and-demand study concludes that recent growth in GP numbers remains insufficient to meet need, with structural drivers (population aging, chronic disease) outpacing capacity [31]. These international experiences echo NL’s situation: stable or slowly growing headcount can coexist with worsening access when outmigration, retirement, scope shifts, administrative burden, and maldistribution depress effective capacity.

Our findings suggest that retention and migration dynamics dominate the near-term trajectory in NL. Even with expected inflows (local FM graduates and IMG licensure), net interprovincial losses, retirements, and scope-shift exits make up significant outflows contributing to the primary care crisis. This points to three pragmatic levers for NL Health Services and partners: (1) Retention first strategies (e.g., team-based redesign that reduces administrative load and supports longitudinal panels) to slow exits and outmigration; (2) Scaleup of Family Care Teams with clear role optimization for nurse practitioners, pharmacists, and other team members to raise effective clinical capacity; and (3) Targeted rural incentives paired with placement supports to counter deepening rural contraction. These directions are consistent with national analyses that highlight team-based care and administrative burden reduction as credible paths to expand access [24, 26].

Our analysis revealed recurring data limitations that reduce precision and comparability. Province-level, up-to-date flows and FTEs are unevenly available, so compiling 2024 inputs (IMG licences, graduate retention, retirements) required piecing together figures from PGME, PRA-NL/CPSNL, NLHS, and NLMA. Definitions also differ: “family physician” may mean direct patient-care providers (CIHI) or a broader licensure group, and “IMG” can refer to training location or a licensure pathway—terms that are not interchangeable. Measurement frames vary (calendar-year headcounts vs. fiscal-year flows; licences vs. billers vs. clinically active), producing residual gaps when comparing flows (2019–2021) with year-to-year stock change. CIHI and NLMA counts do not always align; we therefore used CIHI for time-series consistency and provincial sources to validate and contextualize 2024. Standardization and access remain limited (inconsistent variables/denominators, missing regional breakouts, and unavailable NL-specific indicators), and some data are not collected or not stratified to isolate FPs practising in NL. These constraints call for a provincial, open, standards-based workforce repository linking licensure, payment, and service data.

### Limitations and future research

This study has four main limitations. First, it uses headcount as the primary measure; without complete, comparable FTE data for NL, we approximate effective capacity using a historical FTE-per-head factor, which may under or overstate current effort [22]. Second, CIHI’s entry/exit flow series were publicly available only to 2021; our 2024 reconciliation therefore combines observed historical flows with stakeholder validated adjustments, which may not capture all short-term movements. Third, definitional/timing differences between stock and flow series generate reconciliation residuals, which we document but cannot fully resolve using public data. Fourth, sub-provincial variation is described only where consistently reported; maldistribution may therefore be under-characterized.

Future work should (a) incorporate newer flow years as CIHI releases them; (b) link administrative, licensure, and payment data to estimate true FTE and panel sizes; (c) model policy scenarios (retention gains, IMG pipeline changes, Family Care Team uptake, and role optimization) using system-dynamics or microsimulation; and (d) integrate attachment, ED use for primary care treatable conditions, and equity indicators to tie workforce dynamics to outcomes [7].

## Conclusion

Newfoundland and Labrador’s family-physician supply has been mostly flat but volatile: headcount fell from 680 to 666 between 2014 and 2023, rural representation declined, and net interprovincial losses persisted. CIHI flow data (2019–2021) show substantial turnover. Our result suggests expected entries (MUN graduates, IMGs) did not fully offset exits (migration, retirement, scope shift), yielding ∼658 physicians (∼507 FTE) and about 122 per 100,000 residents. The implication is clear: recruitment alone will not stabilize capacity. Retention-first policies, rapid expansion of team-based care with role optimization, targeted rural supports, and reduced administrative burden are needed, alongside continuous monitoring of flows and patient attachment to guide adjustments.

## Data Availability

The data underlying the results presented in the study are available from CIHI, Statistics Canada, and provincial sources cited in the manuscript.

https://www.mun.ca/medicine/familymedicine/media/production/medicine/family-medicine/DFM_Annual%20Report_2021-2022.pdf

https://www.cihi.ca/en/national-physician-database-metadata

https://www.cihi.ca/en/physicians

https://www.cihi.ca/en/the-state-of-the-health-workforce-in-canada-2023

https://www.cihi.ca/en/taking-the-pulse-measuring-shared-priorities-for-canadian-health-care-2024/better-access-to-primary-care-key-to-improving-health-of-canadians

https://www.mun.ca/medicine/familymedicine/media/production/medicine/family-medicine/2016-Annual-Report.pdf

https://www.mun.ca/medicine/familymedicine/media/production/medicine/family-medicine/DFMAnnualReport_2017.pdf

https://www.mun.ca/medicine/familymedicine/media/production/medicine/family-medicine/DFMAnnualReport_2019.pdf

https://www.mun.ca/medicine/familymedicine/media/production/medicine/family-medicine/DFMAnnualReport_2018-19.pdf

https://www.mun.ca/medicine/familymedicine/media/production/medicine/family-medicine/DFMAnnualReport_201920-FINAL-DOK-SH-JA-Feb-18

## Declarations

## Ethics approval

This study used aggregate, publicly available information and organization-level inputs only; no patient-level or identifiable data were used. Institutional ethics review was therefore not required.

## Consent to participate

Not applicable.

## Data, materials, and/or code availability

All data are publicly available from CIHI, Statistics Canada, and provincial sources cited in the manuscript. Derived tables used in figures are available from the corresponding author upon reasonable request.

## Authors’ contributions

MN conceptualized and designed the study; AM lead the data collection; MN, AM, KAB, and ES contributed to data collection and data analysis. MN drafted the first version of the manuscript. All authors critically revised the manuscript and approved the final version.

